# A novel fusion method of 3D MRI and test results through deep learning for the early detection of Alzheimer’s disease

**DOI:** 10.1101/2024.08.15.24312032

**Authors:** Arman Atalar, Nihat Adar, Savaş Okyay

## Abstract

Alzheimer’s disease (AD) is a prevalent form of dementia that impacts brain cells. Although its likelihood increases with age, there is no transitional period between its stages. In order to enhance diagnostic precision, physicians rely on clinical judgments derived from interpreting health data, considering demographics, clinical history, and laboratory results to detect AD at an early stage. While patient cognitive tests and demographic information are primarily presented in text, brain scan images are presented in graphic formats. Researchers typically use different classifiers for each data format and then merge the classifier outcomes to maximize classification accuracy and utilize all patient-related data for the final decision. However, this approach leads to low performance, diminishing predictive abilities and model effectiveness.

We propose an innovative approach that combines diverse textual health records (HR) with three-dimensional structural magnetic resonance imaging (3D sMRI) to achieve a similar objective in computer-aided diagnosis, utilizing a novel deep learning technique. Health records, encompassing demographic features like age, gender, apolipoprotein gene, and mini-mental state examination score, are fused with 3D sMRI, enabling a graphic-based deep learning strategy for early AD detection. The fusion of data is accomplished by representing textual information as graphic pipes and integrating them into 3D sMRI, a method referred to as the “pipe-laying” method.

Experimental results from over 4000 sMRI scans of 780 patients in the AD Neuroimaging Initiative (ADNI) dataset demonstrate that the pipe-laying method enhances recognition accuracy rates for Early and Late Mild Cognitive Impairment (MCI) patients, accurately classifying all AD patients. In a 4-class AD diagnosis scenario, accuracy improved from 86.87% when only 3D images were used to 90.00% when 3D sMRI and patient health records were included. Thus, the positive impact of combining 3D sMRI with HR on 4-class AD diagnosis was established.

## Introduction

Alzheimer’s disease (AD) is the most common form of dementia, leading to cognitive confusion. The disease advances gradually, resulting in the deterioration of brain cells, impacting memory and cognitive functions, and disrupting daily activities. According to data from the World Health Organization, around 55 million individuals suffer from dementia, with AD constituting 60–70% of these instances. As the elderly population continues to grow, the number of diagnosed patients is projected to escalate to 97 million in the foreseeable future. Developed nations currently observe a 13% prevalence of AD among people aged over 65, and this rate is on the rise (1). Since AD is a type of dementia with a surreptitious onset in the form of episodic memory loss, its early-stage diagnosis poses challenges. Although no definitive cure for AD exists, treatment methods have shown the potential to slow down or even halt disease progression (2). Given that cognitive impairments become noticeable in later stages, rendering effective treatment challenging, early diagnosis becomes crucial to apply interventions that can decelerate or even stop the disease’s advancement.

Mild cognitive impairment (MCI) is the stage between the anticipated age-related decline in memory and cognitive function and the more profound deterioration characteristic of dementia. MCI serves as the intermediary stage and might escalate into dementia as cognitive decline becomes more severe. However, it remains unclear whether the symptoms observed at this stage lead to AD. Investigations centered on sMRI images of patients in this stage have revealed various physical changes in the brain structure. Noteworthy transformations encompass the shrinkage of the hippocampus, pivotal for learning and memory, increased ventricular space, and decreased glucose utilization in certain brain regions. Late and early stages of MCI were established to enhance the comprehension of how MCI impacts Alzheimer’s disease. Early Mild Cognitive Impairment (EMCI) exhibits distinct traits like amyloid buildup, disruptions in functional networks, and variations in brain volume. The ADNI findings indicate that individuals with Late Mild Cognitive Impairment (LMCI) and EMCI face a greater risk of developing dementia linked to Alzheimer’s disease. Contrasted with LMCI patients, those with EMCI demonstrate more heterogeneous characteristics and are more prone to show negative indicators of AD pathology. At the EMCI stage, baseline cognitive function and APOE4 positive status associated with poor cognitive and functional outcomes. In the LMCI phase, the risk of AD escalates with episodic memory impairment (3).

Various learning methods have been developed in this area because of the importance of the early and accurate diagnosis of AD. In addition to imaging modalities, several other factors may be linked to the early diagnosis of AD (2,4). Age, gender, education level, speech pattern, retinal abnormalities, postural kinematic analysis, cerebrospinal biomarkers, neuropsychological measures, and the values of certain genes are essential factors for disease identification (5). In addition, different cognitive and reliable clinical test scores (6), such as the mini-mental state examination, Montreal cognitive assessment, clinical dementia staging score, rey auditory verbal learning test, everyday cognition test, Alzheimer’s disease rating scale, and logical memory test, are required to diagnose AD. After the neurological examination, blood tests, and mental tests, brain imaging should be performed; and in some cases, electroencephalography (EEG), single-photon emission computed tomography (SPECT) lumbar puncture, and psychiatric consultation may be required (2). Using this information together with learning methods will make diagnosis faster and more precise (4).

The majority of deep learning studies have employed neuroimaging techniques, including magnetic resonance imaging (MRI) and positron emission tomography (PET). However, studies utilizing alternative diagnostic data are scarce, and the majority of proposed methods have been designed for use with 2D images. Textual and numerical measurement data, aside from images, are integrated into deep learning approaches for diseases that require early diagnosis. The incorporation of demographic, genetic, and cognitive score data into the diagnostic process contributes to a more precise identification of diseases (5,7).

The diagnostic process for cognitive diseases entails the evaluation of a diverse set of data. Simultaneous and integrated analyses of these data are crucial for early diagnosis. Machine learning-and deep learning-based applications have incorporated data fusion methods to achieve this objective (8). Data fusion involves extracting relevant information from a combination of diverse data originating from various sources. This approach amalgamates multiple data sources to facilitate comprehensive analysis. Data fusion studies involve merging data of the same type into different formats and integrating distinct data types into different formats.

The data fusion methods suggested in the literature for the diagnosis of AD and MCI stages are primarily focused on combining different images, such as MRI and PET (9). Analyzing contextual data such as age, gender, education level, genetics, and cognitive test scores also holds significant importance in the early disease diagnosis. Simultaneously assessing this data alongside images can empower healthcare professionals to achieve early diagnoses.

This study aimed to create a decision support system for physicians, facilitating the differentiation between AD and MCI stages through a simultaneous assessment of images and cognitive test results. To achieve this, textual test results were transformed into 3D pipe image representations and then fused with 3D structural magnetic resonance imaging (3D sMRI) for each patient. This combined approach was named "3D sMRI with Health Records" (3DMRIwHR) for 3D images and "2D sMRI with Health Records” (2DMRIwHR) for 2D images. Given the crucial significance of early AD diagnosis, the proposed 3D convolutional neural networks (CNN) model was employed to classify cognitively normal, early mild cognitive impairment, late mild cognitive impairment, and AD stages in a multi-class fashion.

### Related work

For the early diagnosis of AD, different methods, such as data diversification or model development, are preferred for the multi-classification of its stages. Demographic information, cognitive scores, genetic and neuroimaging data are commonly utilized in data diversification studies. These data are evaluated either as individual features or integrated with the image data.

In a study published in 2018 that focused on separate evaluations of various data, it was observed that the addition of the Mini-Mental State Examination (MMSE) score to MRI improved accuracy in the classification of healthy controls, MCI, and AD groups using different machine learning methods (10). Another study conducted in the same year combined MRI, positron emission tomography (PET), Rey Auditory Verbal Learning Test (RAVLT), Montreal Cognitive Assessment (MoCA), and Electrocochleography Testing (ECogT) standard neuropsychological test scores to classify stages such as AD, EMCI, LMCI, and CN (11).

Data fusion has found application in various neuroimaging studies, with some combining diverse image types while only a minority integrates demographic information with images. An example involves the combination of MRI and PET using a zero-masking strategy, aiming to extract complementary information from distinct data modalities (12). Additionally, Punjabi et al. (13) demonstrated the fusion of MRI and amyloid PET, two widely used imaging modalities. Preprocessing steps for the scans encompassed MRI bias field signal correction, affine recording, and skull stripping. Initially, the methods were individually compared through the utilization of a 3D CNN. In the fusion approach, both imaging datasets underwent parallel processing in separate branches, with features combined in the latter portion of the network through fully connected (FC) layers.

In 2021, a method based on embedded feature selection and fusion using multi-modal neuroimaging was proposed for AD diagnosis. By combining MRI, PET, and Cerebrospinal Fluid (CSF) Factor biomarkers, promising outcomes were obtained for the classification of Healthy Control, MCI, and AD (14). It has been suggested that the performance of the method for multiple classifications of AD can be improved using multi-modal data. In this regard, various modalities have been developed that combine MRI, PET, cerebrospinal fluid (CSF) biomarkers, and genetic features. It has been observed that more effective results were achieved regarding the multi-modal data scored with the linear discriminant analysis method (15).

In addition to multi-class studies, another study was conducted wherein different types of data were integrated into the image, aiming to ascertain the transition from the MCI stage to AD. Through the fusion of different data formats, Pelka et al. (16) employed a long short-term memory-based recurrent neural network (RNN) model for image classification. This data fusion study implies that superior outcomes can be achieved by integrating socio-demographic and medical data for disease diagnosis. Pelka et al. introduced a branding approach wherein socio-demographic and genetic data were encoded using markers on 2D MRI to attain a more advanced image representation and reduce computational load. By employing the ADNI Phase I and Heinz Nixdorf Recall study databases, the study generated five distinct markers denoting age, gender, education, marital status, and ApoE ԑ4 gene values. The outcomes of the study, wherein branded and unbranded images from each marker group underwent separate classification processes, underscored the general impact of branded data on specificity, F1-score, and accuracy performance metrics for both the Heinz Nixdorf Recall and ADNI Phase I databases.

Payan and Montana (17) proposed a two-step approach for feature extraction in a 3D CNN for the classification of sMRI scans. They built a 3D CNN in which the convolutional layers were pretrained using a sparse autoencoder, and directly used 3D sMRI as input data. However, pretraining with the autoencoder was not fine-tuned; therefore, it was suggested that the performance would have been improved by fine-tuning. The model achieved higher accuracy than those of previous studies for multiple classifications. Similarly, a deeply supervised and adaptable 3D CNN (DSA-3D CNN) built on a 3D convolutional autoencoder (3D-CAE) was proposed to capture variations in anatomical sMRI brain scans based on a previous study (18). As a threshold, the 3D-CAE was pretrained using the CAD-Dementia dataset. The proposed model can learn general and transferable features in different regions. The Hierarchical Attention-based Deep Neural Network (HadNet) architecture was suggested for the classification of MRI images that were normalized and skull-stripped (19).

It was concluded that this architecture achieved superior results in terms of sensitivity and specificity. In another study, demographic information and clinical brain activity test scores were marked on MRI images as landmarks for multi-classification. The preprocessed images and landmarks were trained in the CNN model as two separate inputs. During training, the positions of the landmarks were controlled, and the training of landmarks and images was conducted separately. Joint learning was implemented during training. The main deficiency is that landmark detection and landmark-based classification processes are independent (7). The brain images with the landmark skull in the study included colored data on disease classification. Thus, landmarks should be learned independently from CNN in unsupervised learning. In a previous study in which binary and multi-classifications were made, four classes were used: AD, EMCI, LMCI, and NC (20). In this study, 75 samples from each class label were used in the experiments.

It has been revealed that CNN models outperform machine learning methods in the domain of medical image classification. Due to their revolutionary capabilities in capturing multi-region features, CNN are widely used in the medical field to identify different types of diseases using different medical imaging as input data (21). While a majority of data is in 2D format, like chest X-rays or bone X-rays, and is well-suited for 2D CNNs. Advanced medical imaging (3D) produced by sophisticated equipment can be exceptionally well-identified through the utilization of 3D CNNs. Unlike 2D CNNs, 3D CNNs incorporate cross-layer contexts that are not easily discernible through visual inspection. A pivotal distinction between 2D and 3D CNNs lies in the fact that 3D CNNs can retain the spatial information inherent in images. Volumetric CNNs employ 3D filters and generate a 3D volume output by processing the input through a sequence of convolutional layers, including activation, pooling, dropout, and FC layers. Consequently, volumetric CNNs function on the voxels of 3D images while preserving their spatial attributes.

Some studies have analyzed health records in conjunction with 3D image data, and the methodologies they propose involve distinct stages, such as independent feature extraction prior to classification. The autonomy of these stages leads to increased computational complexity and time requirements. Consequently, to address the challenges associated with this approach and achieve early disease diagnosis, this study introduces a method that fuses and simultaneously analyzes textual clinical data and neuroimaging. Moreover, through classification using a deep learning model, this method aims to discern the disease-induced patterns within the brain and identify various stages of MCI and AD. In alignment with this objective, the study also investigates the influence of health records on early diagnosis, assesses the effectiveness of CNNs in analyzing sMRI data, and explores the application of deep learning within the medical domain. The key contributions of this study are outlined as follows.

- Utilizing 3D CNN while preserving the image’s spatial information.
- The 3DMRIwHR fusion method for combining neuroimages and textual data results in reduced algorithm complexity and time requirements.
- Make use of information between layers for early detection of AD using 3D sMRI processing
- Comparative analysis of classification performance between the 3D and 2D CNN models.
- Enhanced early and late MCI diagnosis through the inclusion of HR for both 3D and 2D sMRI classification.

## Material and methods

### Data collection

The data employed in this study were derived from the ADNI dataset. Initiated in October 2004 under the guidance of Dr. Michael W. Weiner, the ADNI dataset comprises clinical, biochemical, and genetic biomarkers, along with neuroimaging modalities such as MRI and PET. These resources can be used for early diagnosis, prevention, and treatment of AD. The study encompassed the initial six years of the ADNI project, denoted as ADNI 1. Subsequent phases include ADNI GO (2009-2011), ADNI 2 (2011-2016), and the ongoing ADNI 3 (2016 to the present). Throughout these stages, image protocols were refined, new participants were incorporated, and diverse data types were integrated into the project.

In this study, a cohort of 780 participants was examined, comprising 178 AD patients, 176 patients with EMCI, 161 patients with LMCI, and 265 CN patients. The 3D sMRI scans were acquired in T1-weighted format, with an in-plane spatial size of 1.25 × 1.25 mm² and a thickness of 1.2 mm in 3D MPRAGE format. In addition to neuroimaging data, the study encompassed demographics, cognitive metrics, and genetic information of the participants. Patient health records, encompassing demographic factors like age and gender, ApoE-ε4 genetic data, and MMSE cognitive measure data were fused with 3D sMRI data. Consequently, a 3D CNN model based on 3DMRIwHR images was deployed to classify the four distinct cognitive states associated with dementia: normal cognition, early mild cognitive impairment, late mild cognitive impairment, and AD. Table 1 provides details about the participant count and other pertinent information for each group. The majority of participants were over 65 years old, with MMSE scores exceeding 19, and a greater prevalence of negative ApoE-ε4 values compared to positive ones.

**Table 1.**
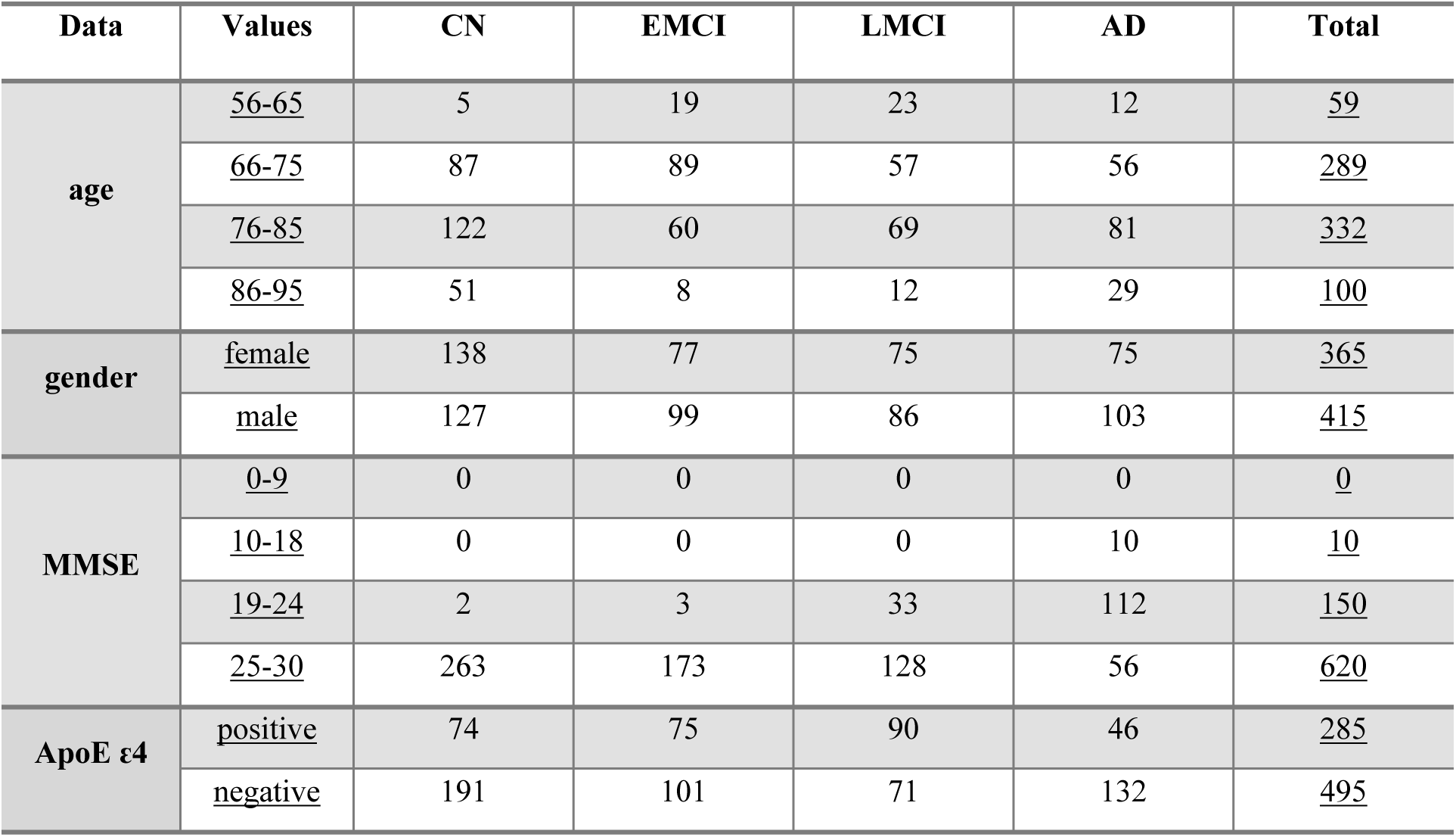
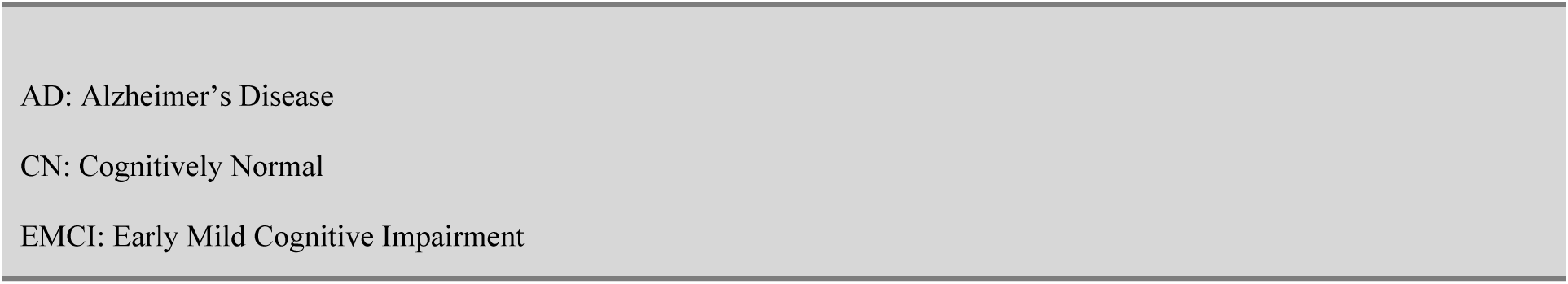
Demographic, Cognitive Score and Gene Data of All Subjects.

| Data | Values | CN | EMCI | LMCI | AD | Total |
| --- | --- | --- | --- | --- | --- | --- |
| age | <u>56-65</u> | 5 | 19 | 23 | 12 | <u>59</u> |
|  | <u>66-75</u> | 87 | 89 | 57 | 56 | <u>289</u> |
|  | <u>76-85</u> | 122 | 60 | 69 | 81 | <u>332</u> |
|  | <u>86-95</u> | 51 | 8 | 12 | 29 | <u>100</u> |
| gender | <u>female</u> | 138 | 77 | 75 | 75 | <u>365</u> |
|  | <u>male</u> | 127 | 99 | 86 | 103 | <u>415</u> |
| MMSE | <u>0-9</u> | 0 | 0 | 0 | 0 | <u>0</u> |
|  | <u>10-18</u> | 0 | 0 | 0 | 10 | <u>10</u> |
|  | <u>19-24</u> | 2 | 3 | 33 | 112 | <u>150</u> |
|  | <u>25-30</u> | 263 | 173 | 128 | 56 | <u>620</u> |
| ApoE ε4 | <u>positive</u> | 74 | 75 | 90 | 46 | <u>285</u> |
|  | <u>negative</u> | 191 | 101 | 71 | 132 | <u>495</u> |

AD: Alzheimer's Disease
CN: Cognitively Normal
EMCI: Early Mild Cognitive Impairment

### Preprocessing

Taking computational efficiency into consideration, the initial step involved preprocessing the images through the utilization of FMRIB Software Library v6.0.5.2. The 3D sMRI data, initially registered using FMRIB’s Linear Image Registration Tool module, underwent bias field correction using the FLIRT module. This procedure eliminated the density gradient that could impact the segmentation algorithm. MRI measurements pertaining to the skull, skin, fat, muscle, neck, and eyeballs were deemed insignificant for disease identification. A brain extraction tool module was applied to remove non-brain tissues, including the skull. Subsequently, intensity normalization was executed to rectify density variations potentially arising due to differing image sizes. The final preprocessing stage involved resizing the 3D images from 176 × 240 × 256 to 50 × 30 × 20, a resolution validated in [20]. The adoption of larger image sizes would result in heightened computational complexity, rendering them unfeasible for implementation on conventional computers.

To compare the impact of fusing health records with MRI scans for multi-class image classification, both 2D and 3D image data fusion methodologies were employed. From the preprocessed and unresized 3D images, 2D slices were extracted after skull-stripping. In the final phase of 2D MRI preprocessing, these slices were saved in PNG format with 256 × 256 pixels. Within the ADNI dataset, subjects with health records encompassing age, gender, ApoE-ε4, and MMSE are limited and are imbalanced across AD, LMCI, EMCI, and CN categories. Following a 20% allocation for testing, data augmentation was undertaken to increase the dataset’s size for both the 2D and 3D CNN models. The number of images per class, initially set at 1000, was doubled through this process. Consequently, the shuffled dataset, encompassing 8000 MRI scans, was subsequently partitioned into two sets: training and validation.

### Fusing 3D sMRI with Health Records: 3DMRIwHR Method

In the diagnosis of Alzheimer’s disease, a specialist physician conducts a range of examinations after reviewing the patient’s medical history. These examinations encompass neuropsychological, neurophysiological, genetic, laboratory, neuroimaging, and nuclear medicine tests. Disease diagnosis isn’t solely reliant on neuroimaging; it also involves the analysis of diverse data types. Healthcare professionals face restricted opportunities to effectively utilize these varied data forms. Despite the growing body of research on this subject, MRI remains the preferred approach. In this study, we introduced a method named 3DMRIwHR, which facilitates the assessment of neuroimaging data by combining demographic, genetic, and cognitive test score outcomes. When selecting test results represented by 3D-HR markers, factors that heightened the risk of AD symptoms were considered.

3D sMRI contains additional information between layers, a feature not present in 2D sMRI. However, extracting this information through visual inspection of 3D sMRI images is not trivial. Machine learning techniques, however, can effectively harness this information. To underscore the interdimensional impact of the 3D method, a fusion procedure was applied to 2D slices derived from 3D images within the proposed deep learning model. This approach, utilizing the same data, is designated as 2DMRIwHR.

The approach used in the study encompassed the fusion of text-based information with sMRI scans through the utilization of 3D volumetric pipe-shaped markers. In marker design, preference was given to half and full rectangular prism shapes in black or white, with each color symbolizing specific attributes. These textual attributes included age, gender, ApoE-ԑ4 gene value, and MMSE score ranges. Figure 1 illustrates the attribute values associated with these colored volumetric pipes. The characteristics of the markers utilized in the fusion of 2D images, along with their corresponding value ranges and arrangement on the image, were applied similarly to the 3DMRIwHR method, with no changes except for spatial criteria. Upon extracting the 2D images as detailed in the data preprocessing section, they were combined with 2D markers that represented textual information. This methodology was labeled as 2DMRIwHR. Following this technique’s application, training was done on the proposed 2D model without including any additional image enhancements

**Figure 1.**
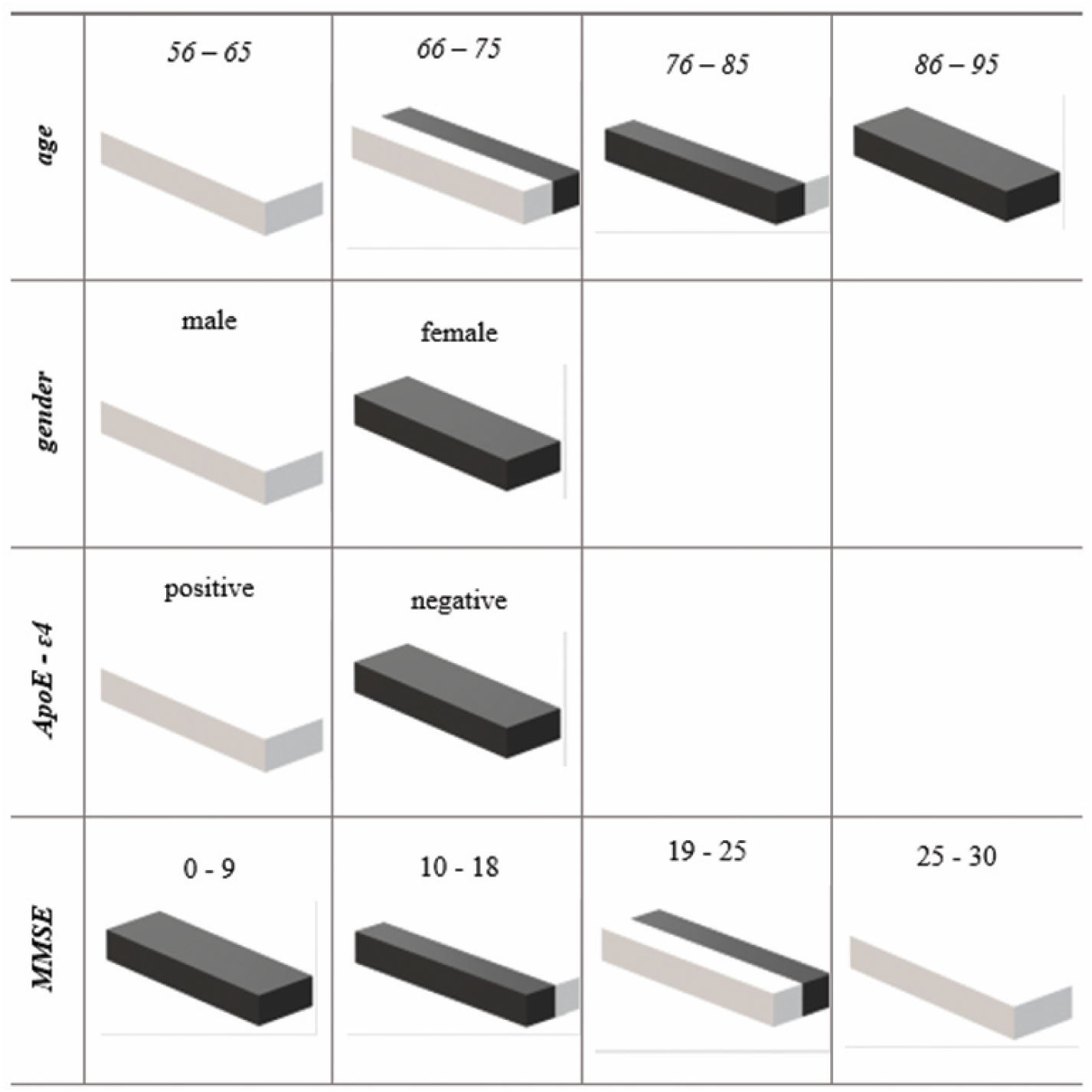
3D pipe shape coding for health records.

In this study, four distinct health records — age, gender, MMSE, and ApoE-ԑ4 gene data — were utilized to construct 3D-HR pipes, as shown in Figure 1. Each of the 3D pipes created using the 3DMRIwHR method had a height of eight pixels, a width of 20 pixels, and a depth of 256 pixels.

The processing involved 3D images in NIfTI format, with dimensions of 176 × 240 × 256, carried out in the axial plane, spanning from pixel points (2, 10) to (10,49). Gaps of size (5 × 4) were deliberately left between the pipe markers to ensure that volumetric markers did not generate any erroneous correlations. 3D-HR pipes were located in the corners of the 3D sMRI scans for the purpose of data fusion. The corner’s selection was such that it didn’t impede the 3D brain image. In the case of the 2DMRIwHR method, 2D planar markers, with dimensions of eight pixels in height and 20 pixels in width, were employed. The positioning process on the image closely resembled that of the 3DMRIwHR method.

To visualize the Nifti scans, the 3D axial preprocessed and 3DMRIwHR images were transformed into PNG files, as shown in Figure 2. The 3D-HR pipes representing age, gender, ApoE-ԑ4, and MMSE scores on the image were placed in a specific order.

**Figure 2.**
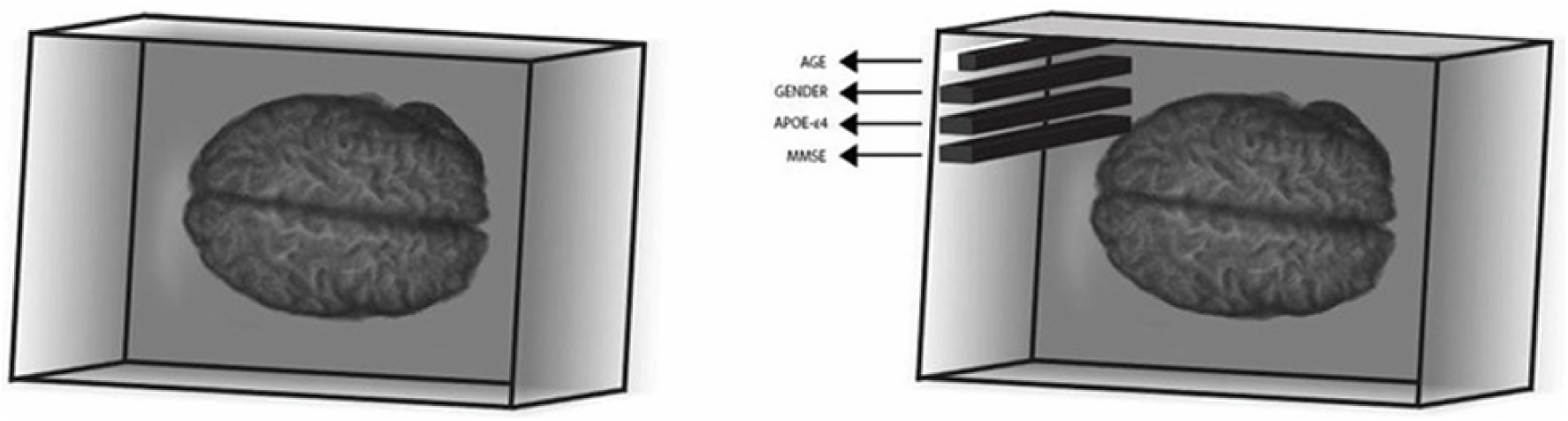
3D MRI with and without 3D-HR pipes.

2D MRI images were extracted from the 3D sMRI images by means of slicing. After the slicing phase, the 2D images in Nifti format were combined with health records using 2D encoding. The markers employed in the 2D images denoted the same values as those in the 3D-HR pipes. While the alignment of marker images remained consistent, due to the 2D and planar nature of the images, the 2D markers were depicted as rectangles, measuring eight pixels in height and 20 pixels in width.

### Classification for Decision Support

CNNs are a specific type of neural network designed for processing data organized in matrix form. Convolutional networks have proven effective in real-world applications, yielding satisfactory outcomes not only with large datasets but also when adeptly modeled for smaller datasets. In the realm of image processing studies [19,20,22], the architecture of CNN best fits the concept of learning regional patterns by transforming pixel data into matrices. Given that dementia impacts specific brain regions, we employed 2D and 3D CNN architectures to classify the four classes, namely CN, EMCI, LMCI, and AD.

### 3D CNN method

The proposed model comprises three 3D convolutional layers and three 3D max pools. Following these layers, a dropout layer was introduced, succeeded by a flattened layer. At the model’s last layer, there existed an FC layer employing a softmax activation function. The 3D convolutional layers were all equipped with 3 × 3 × 3 kernels and 1 × 1 × 1 stride values, featuring 64 filters in the initial two convolutional layers and 128 filters in the final layer. The pooling layer kernels were all of size 3 × 3 × 3, with stride values set at 2 × 2 × 2. The number of training parameters was 1,710,660, whereas the number of non-trainable parameters was 0. For optimization during parameter training, the adaptive moment-estimation (ADAM) optimization algorithm was utilized with a learning rate of 0.001.

Since the rectified linear unit (RELU) is currently the most commonly applied function [21], it was used as the activation function in the three convolutional layers. A dropout value of 0.5 was selected to mitigate the risk of overfitting. Following the softmax activation function in the final FC layer, a 4-class disease classification was executed. The architectural layout of the 3D CNN model can be observed in Figure 3.

**Figure 3.**
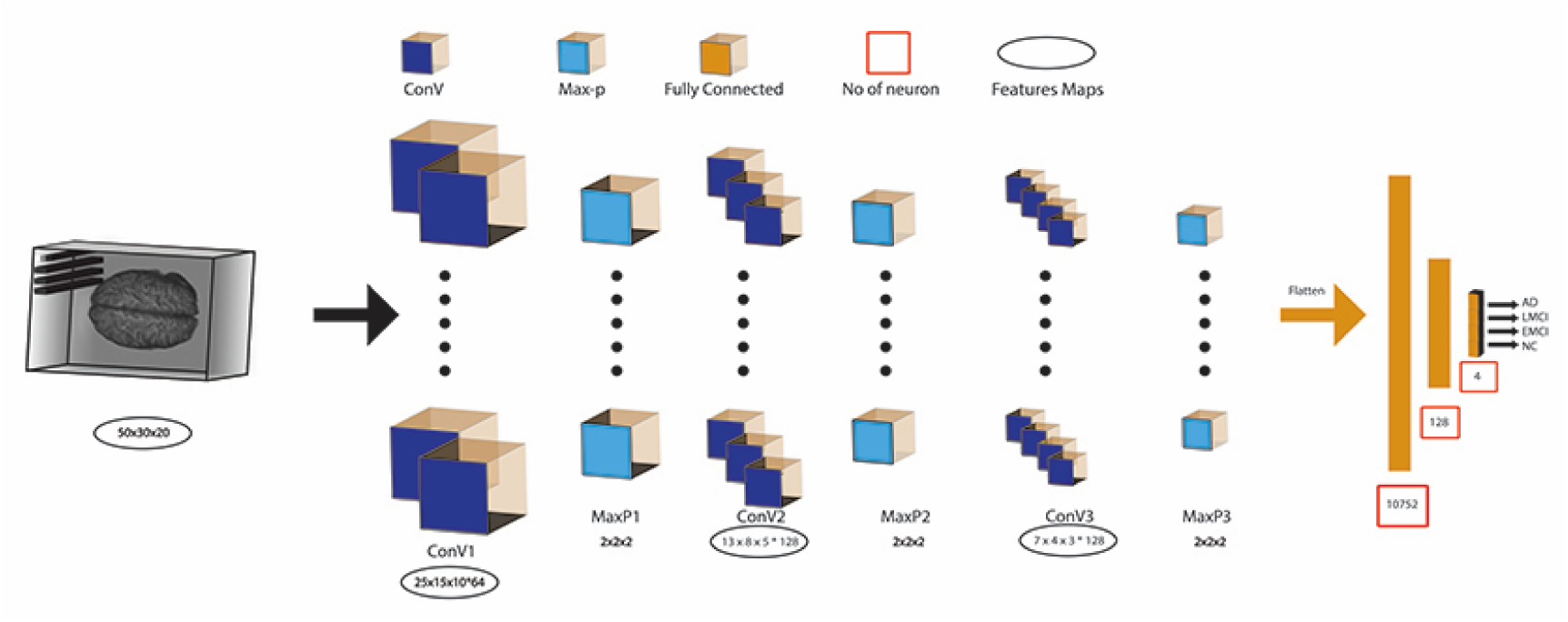
3D CNN Model Architecture.

### 2D CNN Method

In the 2D CNN model, there were three 2D convolutional layers accompanied by three 2D max pools. These were followed by a dropout layer, succeeded by a flattened layer. The model’s conclusion included an FC layer employing a softmax activation function. The 2D convolutional layers all consisted of 3 × 3 kernels with 1 × 1 stride values, with 64 filters present in the initial two convolutional layers and 128 in the last convolutional layer. The pooling layer kernels were all 3 × 3 in size, with stride values set at 2 × 2. The number of training parameters was 16,889,284, whereas the number of non-trainable parameters was 0. An optimization algorithm, specifically "ADAM" was utilized with a learning rate of 0.001 during parameter training.

As for the activation function, RELU was utilized for the three convolutional layers due to its widespread application [21]. A dropout value of 0.5 was selected to counteract overfitting. The classification of the four dementia groups was carried out based on the softmax activation function in the final FC layer.

### Experimental Design

In our study, the proposed models considered the impact of health records on classifier performance. To illustrate the effect of health records on recognition rates, the comparison was made between 2D MR vs. 2DMRIwHR and 3D MR vs. 3DMRIwHR, utilizing 2D CNN and 3D CNN deep-learning techniques, respectively.

The 2D and 3D classification methods were implemented using the Keras 2.8.0 library in Python 3.9, built on TensorFlow-gpu 2.8.0. The execution occurred on a PC equipped with an NVIDIA RTX2060 GPU running the Windows operating system. In the network, the Adaptive Moment Estimation (ADAM) optimizer was employed, starting with an initial learning rate of 0.001. A dropout rate of 0.5 was set to mitigate overfitting. For 3D classification, the batch size was set to 64, while the 2D classification employed a batch size of 128.

The image count for each class was divided into two sets using a 0.8 training and 0.2 testing ratio. Subsequently, data augmentation was applied to the train set, effectively doubling the number of images in each class. As a result, the overall training dataset containing 8000 images was divided into a training set (comprising 64% of the images), a validation set (consisting of 16% of the images). During the classification phase, only images belonging to the class with the lowest image count were utilized to accurately assess the impact of the pipe-laying process. This approach ensured balanced representation among classes.

### Data Augmentation

The image count for each class was divided into two sets using a 0.8 training and 0.2 testing ratio. Subsequently, data augmentation was applied to the train set, effectively doubling the number of images in each class. As a result, the overall training dataset containing 8000 images was divided into a training set (comprising 64% of the images), a validation set (consisting of 16% of the images).

The 2D and 3D classification methods were implemented using the Keras 2.8.0 library in Python 3.9, built on TensorFlow-gpu 2.8.0. The execution occurred on a PC equipped with an NVIDIA RTX2060 GPU running the Windows operating system. In the network, the Adaptive Moment Estimation (ADAM) optimizer was employed, starting with an initial learning rate of 0.001. A dropout rate of 0.5 was set to mitigate overfitting. For 3D classification, the batch size was set to 64, while the 2D classification employed a batch size of 128.

## Results and Discussion

The 3D CNN method using only MRI achieved an accuracy of 86.87%, while the inclusion of health records (3DMRIwHR) improved accuracy by 3.13% for multi-class classification of preprocessed medical images involving AD, LMCI, EMCI, and CN categories. Similarly, the 2D CNN method without health records achieved an accuracy of 81.43%, which increased to 87.00% when health records were integrated. Given the improved performance of both 2D and 3D CNN models with health records, it can be inferred that combining health records with MRI data leads to superior outcomes. Furthermore, it was observed that the 3D CNN model with health records outperformed the 2D CNN model with health records due to its ability to leverage spatial information between the 2D layers of MRI. As depicted in Figures 4 and 5, the training and validation accuracies provide evidence that the proposed models function effectively without overfitting.

**Figure 4.**
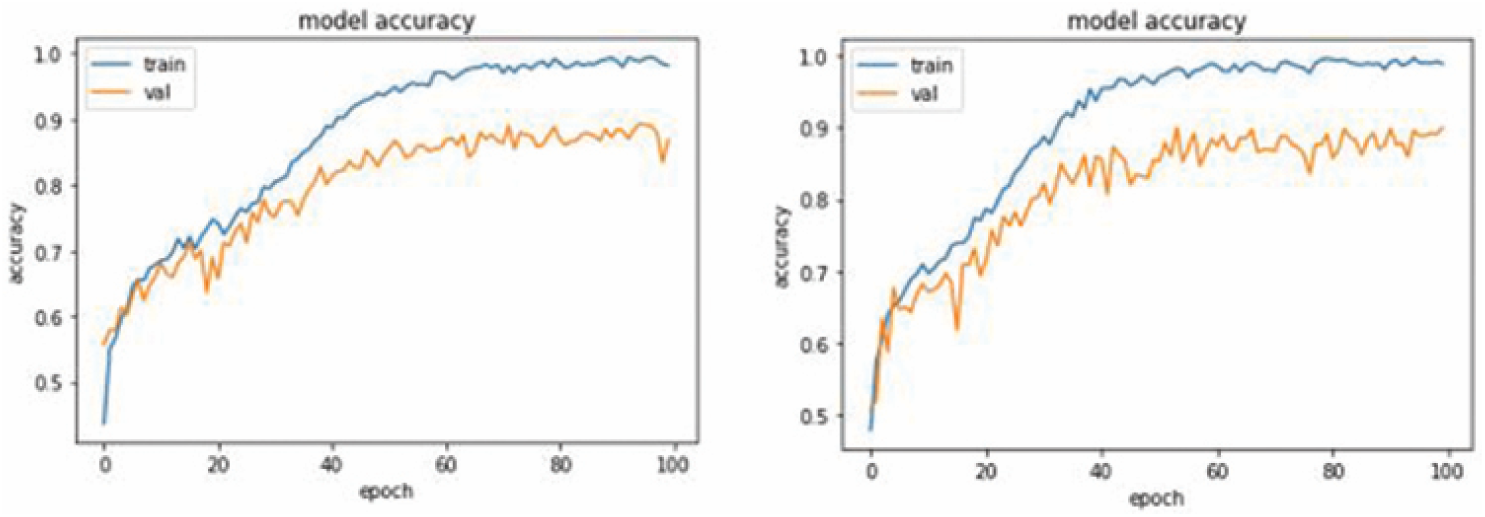
Training and validation accuracy and loss for 3D classification model.

**Figure 5.**
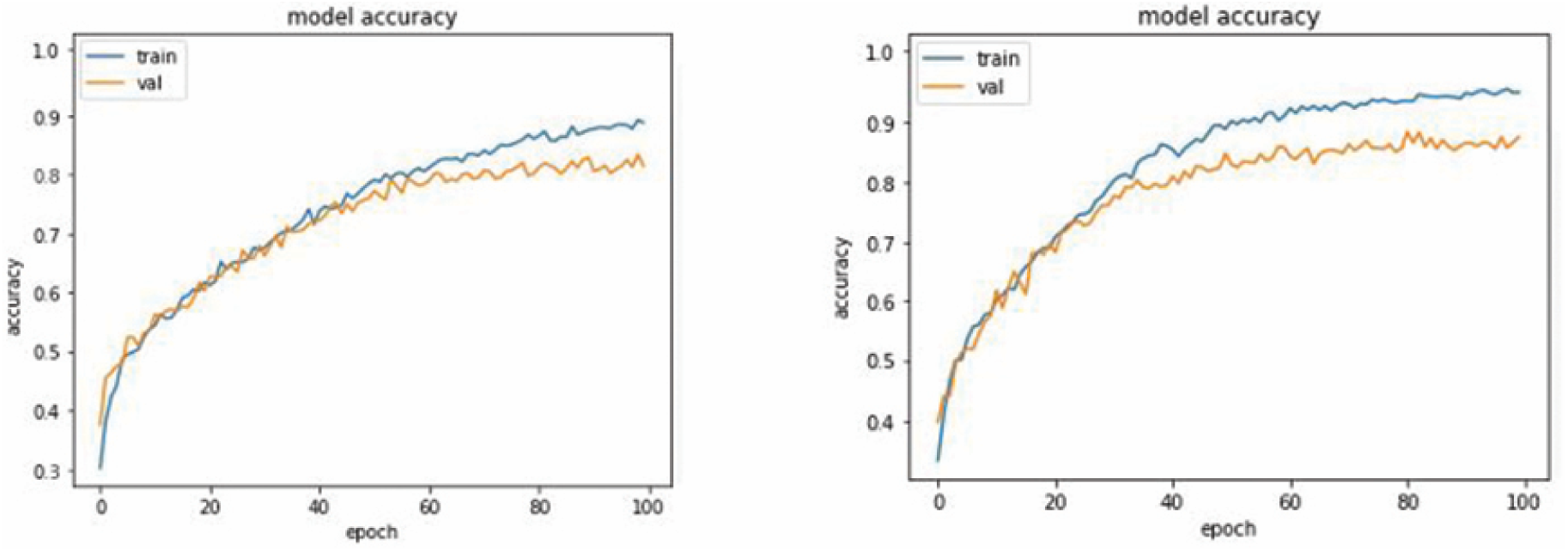
Training and validation accuracy and loss for 2D classification model (a) with and (b) without 3D-HR data.

Evidently, the 3D CNN model surpassed the 2D CNN model in classifying both pre-and post-method datasets. The utilization of information across layers in 3D sMRI scans, as opposed to 2D images, grants the 3D CNN model a greater classification advantage. Illustrated in Figures 4 and 5, the difference in accuracy rates before and after applying the method was less pronounced in 3D classification compared to 2D classification, mainly due to the richer features present in 3D images. Notably, the additional features introduced alongside health records exhibited a more substantial impact on the classification accuracy of pre-and post-method 2D images, despite their lower overall feature count.

The confusion matrix illustrates the performance of the proposed 3D classifier in distinguishing various class labels when the 3DMRIwHR method is not utilized, as depicted in Figure 6. Without the application of the 3DMRIwHR method, 691 out of 800 images were accurately predicted, while 109 were predicted incorrectly. In the post-method classification, 720 out of 800 images were correctly predicted, and 80 were inaccurately predicted. By ranking disease stages by severity as AD, LMCI, EMCI, and CN, the error matrix revealed that the detection capability improved with the 3DMRIwHR method as the disease progressed to more severe stages.

**Figure 6.**
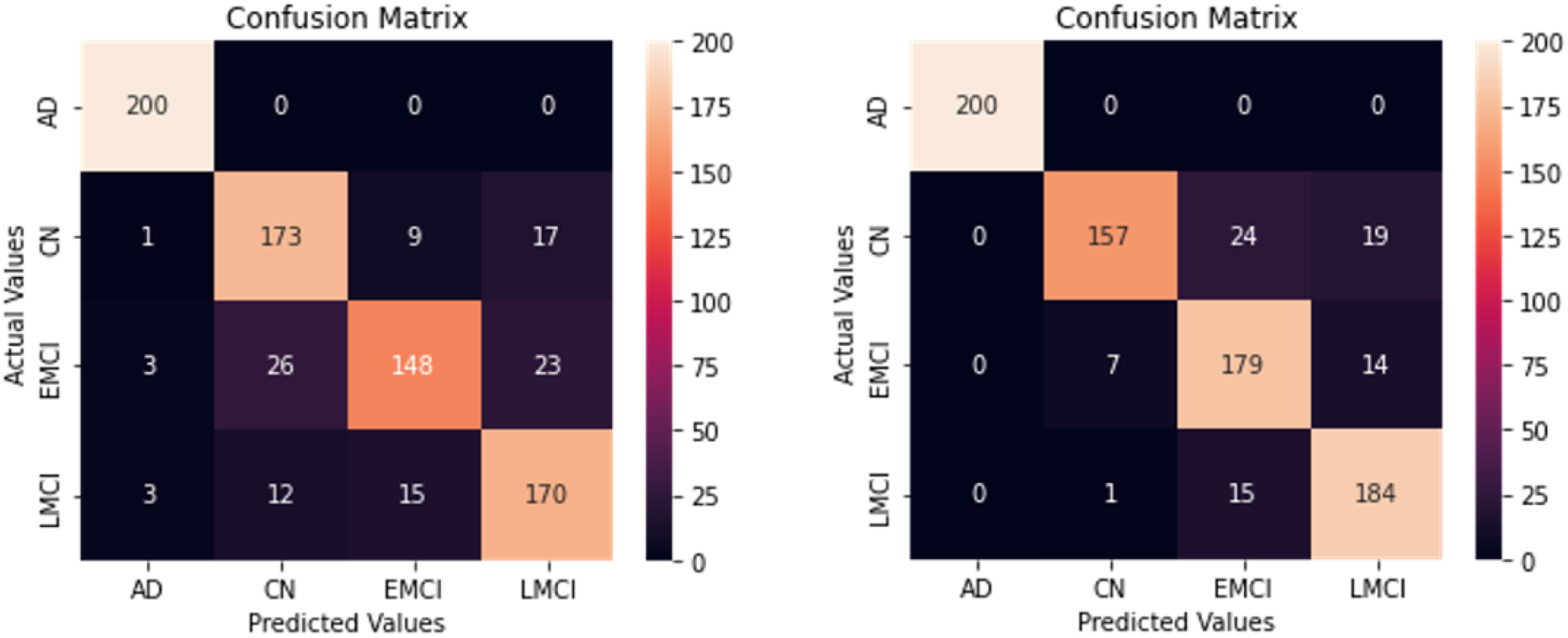
The confusion matrix for preprocessed and pipe layed data 3D classification model.

The confusion matrix shows the effectiveness of the proposed 2D classifier in categorizing different class labels when the 2DMRIwHR method is both applied and not applied, as shown in Figure 7. In cases where the 2DMRIwHR method was not used, 645 of the 800 images were accurately predicted, while 155 were incorrectly predicted. Following the post-method classification, 698 out of 800 images were correctly predicted, and 102 were predicted inaccurately. Similar to the 3D classifier, a pattern emerged in the confusion matrix for the 2D classifier, indicating improved detection ability with the 2DMRIwHR method as the disease severity escalated.

**Figure 7.**
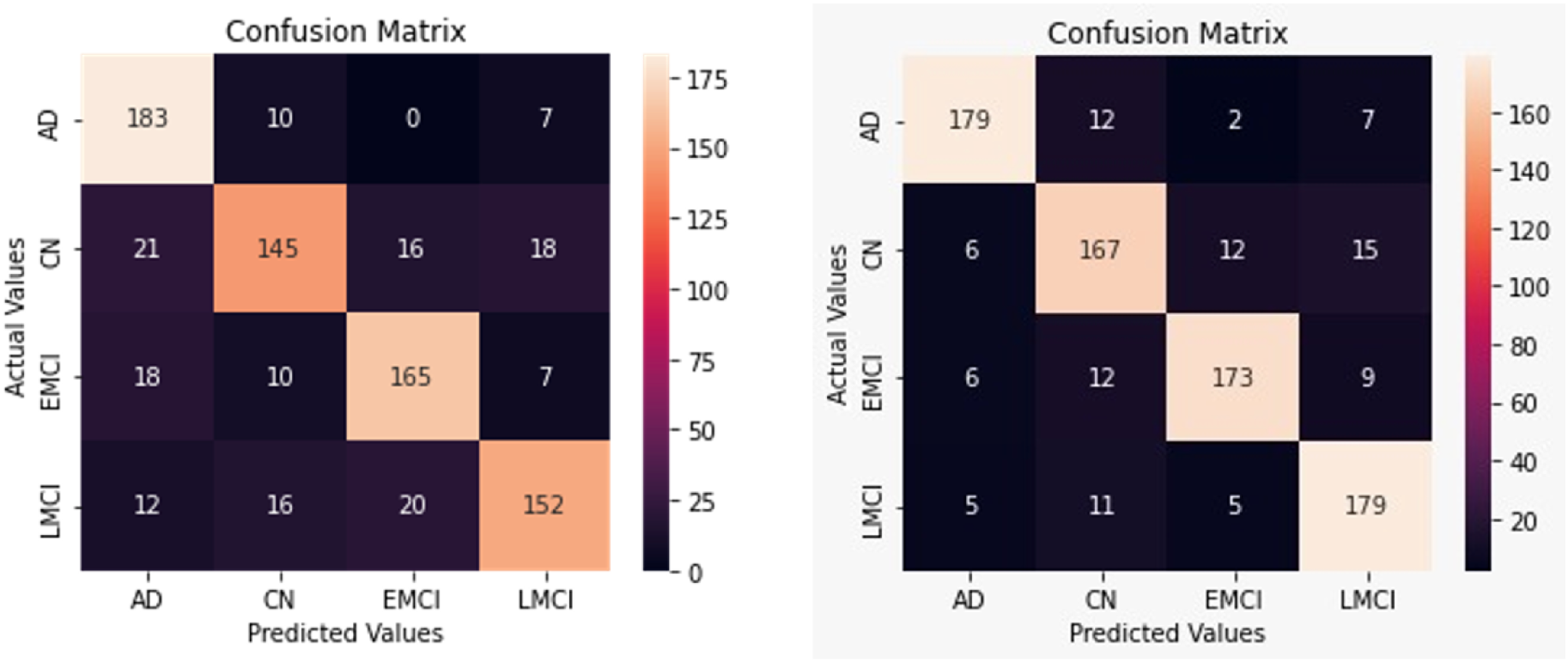
The confusion matrix for preprocessed and pipe layed data 2D classification model.

Comparing results before and after the application of the 3DMRIwHR method in the order of class levels CN, EMCI, LMCI, and AD revealed an increase in the predictive capability of the proposed model with rising disease levels. Notably, the 3D CNN model accurately identified all images in the AD class, regardless of the presence of this method. The highest estimation for the healthy class (CN) was achieved when the 3DMRIwHR method was applied, utilizing a single MMSE 3D-HR marker. Within the EMCI class, the highest estimation occurred in the classification following the 3DMRIwHR method, wherein all markers were applied collectively. The proposed method notably heightened predictive accuracy for the early MCI level, which signifies the disease’s initial stage and aligns with the study’s goal of early diagnosis.

Regarding the LMCI class, the method’s application using a single age 3D-HR marker resulted in the highest predictive accuracy. The decreased classification accuracy observed when combining images with ApoE ε4 3D-PI markers, compared to the rate without the method, can be attributed to the dataset’s composition: 285 participants exhibited positive values, while 495 displayed negative values.

The evaluation of the model encompassed the analysis of accuracy, loss, confusion metrics, F1 score, recall, and precision parameters. Table 2 presents the performance metrics of the model after 100 learning epochs.

**Table 1.** Comparison of the Performance Metrics of 3D CNN Model with and without 3DMRIwHR.

|  | 3D MRI |  |  | 3DMRIwHR |  |  |  |
| --- | --- | --- | --- | --- | --- | --- | --- |
|  | Precision | Recall | F<sub>1</sub>-Score | Precision | Recall | F<sub>1</sub>-Score | Support |
| **AD** | 0.97 | 1.00 | 0.98 | 1.00 (+0.03) | 1.00 | 1.00 (+0.02) | 200 |
| **LMCI** | 0.81 | 0.85 | 0.83 | 0.85 (+0.04) | 0.92 (+0.7) | 0.88 (+0.05) | 200 |
| **EMCI** | 0.86 | 0.74 | 0.80 | 0.82 (-0.04) | 0.90 (+0.16) | 0.86 (+0.04) | 200 |
| **CN** | 0.82 | 0.86 | 0.84 | 0.95 (+0.13) | 0.79 (-0.7) | 0.86 (+0.02) | 200 |
| **Avg** | 0.86 | 0.86 | 0.86 | 0.91 (+0.05) | 0.90 (+0.04) | 0.90 (+0.04) | 800 |

The precision column displays values signifying the actual criteria for accurately classified images among all samples categorized as positive. Without utilizing this method, the precision criterion for correctly classifying patients with AD was 0.97. With the implementation of this method, this ratio improved to 1.00. Notably, precision values exhibit a progressive increase in accordance with the proposed method as patient groups are arranged from worse to better.

The second column presents the sensitivity (recall) scale, which signifies the model’s capability to detect positive samples. For the patients with EMCI, the sensitivity rate elevated from 0.74 to 0.90. Application of the 3DMRIwHR method led to increased sensitivity rates in diagnosing patient groups but decreased rates in the healthy group.

The F1 score, a harmonic mean of precision and sensitivity values, was chosen to avoid incorrect model selection in unbalanced datasets. Given the study’s use of balanced datasets, the F1 score closely mirrored the precision values in the research outcomes. Noticeably, these values escalated for all classes, with no imbalance between precision and sensitivity. While F1 scores did not decrease for cases before and after applying the method, enhancements were evident in the AD, LMCI, and CN classes.

For the 2D CNN performance metrics, Table 3 details precision, sensitivity, and F1 scores for each class in scenarios with and without the proposed method. Prior to employing the 2DMRIwHR method, the precision criterion for AH images was 0.78, which increased to 0.86 with the method. The sensitivity also grew by 0.10 for patients with EMCI. F1 scoring values witnessed elevation for all classes and there was no imbalance between the precision and sensitivity values. In post-method classification, all metrics in the model exhibited improvement, barring the sensitivity value for the CN class.

**Table 2.**
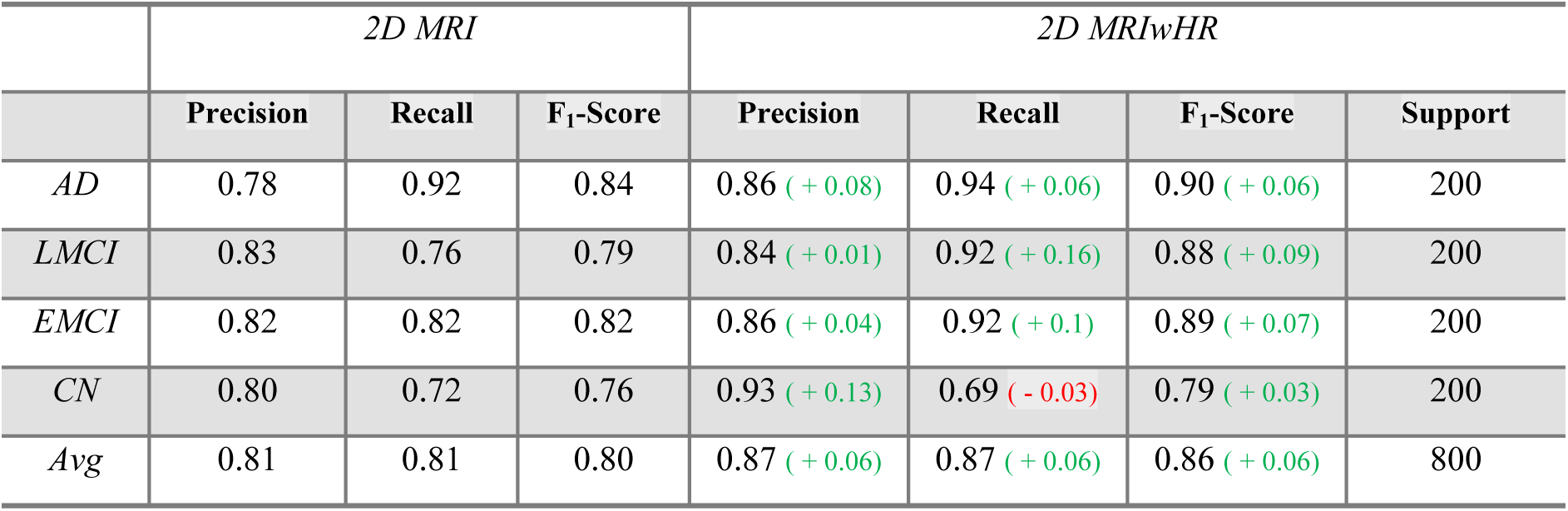
Comparison of the Performance Metrics of 2D CNN with and without 2DMRIwHR.

|  | *2D MRI* |  |  | *2D MRIwHR* |  |  |  |
| --- | --- | --- | --- | --- | --- | --- | --- |
|  | **Precision** | **Recall** | **F<sub>1</sub>-Score** | **Precision** | **Recall** | **F<sub>1</sub>-Score** | **Support** |
| *AD* | 0.78 | 0.92 | 0.84 | 0.86 (+ 0.08) | 0.94 (+ 0.06) | 0.90 (+ 0.06) | 200 |
| *LMCI* | 0.83 | 0.76 | 0.79 | 0.84 (+ 0.01) | 0.92 (+ 0.16) | 0.88 (+ 0.09) | 200 |
| *EMCI* | 0.82 | 0.82 | 0.82 | 0.86 (+ 0.04) | 0.92 (+ 0.1) | 0.89 (+ 0.07) | 200 |
| *CN* | 0.80 | 0.72 | 0.76 | 0.93 (+ 0.13) | 0.69 (- 0.03) | 0.79 (+ 0.03) | 200 |
| *Avg* | 0.81 | 0.81 | 0.80 | 0.87 (+ 0.06) | 0.87 (+ 0.06) | 0.86 (+ 0.06) | 800 |

Separate processes were conducted on both 2D and 3D images to assess the impact of the data fusion method across dimensions. Based on performance metrics, classification outcomes for 3D-HR data were consistently superior to those achieved without 3D-HR data in both models. Moreover, our study affirmed that a 3D CNN outperformed a 2D CNN in MRI classification accuracy. Similarly, akin to the 3D fusion method, patient health records were incorporated with 2D MRI images. However, extracting meaningful information from 3D sMRI visually is a nontrivial task. It has been evidenced that deep learning techniques applied to 3D sMRI, leveraging inter-layer information, yielded enhanced recognition results compared to other techniques employing 2D sMRI.

This data fusion study encompasses several factors contributing to its uniqueness, such as preprocessing and 3D-HR encoding, simultaneous utilization of neuroimages and health records, and harnessing the 3D CNN architecture to achieve elevated accuracy through learning regional patterns, distinguishing it from prior studies. To highlight the impact of the fusion method, results both with and without the fusion method are presented. We trained markers as features, transforming them into features during the preprocessing stage, thereby avoiding separate training and reducing computational time.

## Conclusion

Computer-aided systems have the potential to aid doctors in achieving faster and more precise diagnoses during the early stages of AD. To achieve this goal, data beyond patient images is employed within the diagnostic process. Consequently, the types of data utilized can be diverse. CAD systems can experience enhancements when incorporating various data types. In our study, alongside image data, demographic information like age, gender, MMSE, and ApoE-ε4 was integrated.

In this study, classification was performed using a CNN architecture based on fused sMRI data, with textual health records coded as image markers to detect AD. Data fusion was performed on the preprocessed MRI by transforming participant attributes such as demographic features like age, gender, ApoE-ε4 gene information, and MMSE cognitive scores into image markers. In multi-class classification, accuracy values were obtained for the 2D and 3D CNN. The test results demonstrated that fusing health records increased the recognition rates for Alzheimer’s diagnosis. Our study revealed that different types of data can be used to detect AD. Adding health records to MRI scans increases the AD diagnosis recognition rates. The accuracy of 3DMRIwHR is better than that of 2DMRIwHR because the 3D image architecture exploits the spatial information between 2D slices in sMRI. However, the 2D CNN classification method produces a better accuracy than the 2D LSTM-based RNN because of its pattern detection approach. The 3DMRIwHR method achieved a high recognition accuracy with multi-class dementia evolution. Thus, practitioners can gain better insights by observing the early phases of dementia diagnosis based on multiple features.

Reducing examination time and facilitating diagnoses through CAD systems can expedite the dementia diagnosis process for healthcare professionals. Our study demonstrates that diverse data formats can be combined to achieve enhanced accuracy rates while maintaining lower computational complexity. This empowers doctors and specialists to diagnose dementia by utilizing all available patient-related data.

In conclusion, our study proposes that the 3D data fusion approach can serve not only for classification but also for predicting convertible MCI.

## Data Availability

The data is available at http://adni.loni.usc.edu/data-samples/access-data. Data collection and sharing for this project was provided by the Alzheimer”s Disease Neuroimaging Initiative (ADNI) (National Institutes of Health Grant U01 AG024904) and DOD ADNI (Department of Defense award number W81XWH-12-2-0012). ADNI is funded by the National Institute on Aging, the National Institute of Biomedical Imaging and Bioengineering, and through generous contributions from the following: AbbVie, Alzheimer”s Association Alzheimer”s Drug Discovery Foundation Araclon Biotech BioClinica, Inc. Biogen Bristol-Myers Squibb Company CereSpir, Inc. Cogstate Eisai Inc. Elan Pharmaceuticals, Inc. Eli Lilly and Company EuroImmun F. Hoffmann-La Roche Ltd and its affiliated company Genentech, Inc. Fujirebio GE Healthcare IXICO Ltd. Janssen Alzheimer Immunotherapy Research & Development, LLC. Johnson & Johnson Pharmaceutical Research & Development LLC. Lumosity Lundbeck Merck & Co., Inc. Meso Scale Diagnostics, LLC. NeuroRx Research Neurotrack Technologies Novartis Pharmaceuticals Corporation Pfizer Inc. Piramal Imaging Servier Takeda Pharmaceutical Company and Transition Therapeutics. The Canadian Institutes of Health Research is providing funds to support ADNI clinical sites in Canada. Private sector contributions are facilitated by the Foundation for the National Institutes of Health (www.fnih.org). The grantee organization is the Northern California Institute for Research and Education, and the study is coordinated by the Alzheimer”s Therapeutic Research Institute at the University of Southern California. ADNI data are disseminated by the Laboratory for Neuro Imaging at the University of Southern California.

https://adni.loni.usc.edu/

## Additional Information and Declarations

### Author Contributions

- Arman Atalar conceived and designed the experiments, performed the experiments, analyzed the data, prepared figures and/or tables, authored or reviewed drafts of the paper, performed the computation work, and approved the final draft.
- Savas Okyay conceived and designed the experiments, analyzed the data, authored or reviewed drafts of the paper, and approved the final draft.
- Nihat Adar conceived and designed the experiments, analyzed the data, authored or reviewed drafts of the paper, and approved the final draft.

## Acknowledgements

We thank neurologist Dr. Serdar Eren (LÖSANTE) for providing us with information about the clinical procedures.

## Human Ethics

The following information was supplied relating to ethical approvals (i.e., approving body and any reference numbers):

Alzheimer’s Disease Neuroimaging Initiative: “All ADNI data are shared without embargo through the LONI Image and Data Archive (IDA), a secure research data repository. Interested scientists may obtain access to ADNI imaging, clinical, genomic, and biomarker data for the purposes of scientific investigation, teaching, or planning clinical research studies. Access is contingent on adherence to the ADNI Data Use Agreement and the publications’ policies outlined in the documents listed below. Note: documents are subject to updates by ADNI.” More information is available at https://adni.loni.usc.edu/data-samples/access-data/.

## Data Availability

The following information was supplied regarding data availability:

The project repository containing the supplementary material and other sources are available at GitHub: https://github.com/armanatalar/3DMRIwHR.

The data is available at http://adni.loni.usc.edu/data-samples/access-data/.

Data collection and sharing for this project was provided by the Alzheimer’s Disease Neuroimaging Initiative (ADNI) (National Institutes of Health Grant U01 AG024904) and DOD ADNI (Department of Defense award number W81XWH-12-2-0012). ADNI is funded by the National Institute on Aging, the National Institute of Biomedical Imaging and Bioengineering, and through generous contributions from the following: AbbVie, Alzheimer’s Association; Alzheimer’s Drug Discovery Foundation; Araclon Biotech; BioClinica, Inc.; Biogen; Bristol-Myers Squibb Company; CereSpir, Inc.; Cogstate; Eisai Inc.; Elan Pharmaceuticals, Inc.; Eli Lilly and Company; EuroImmun; F. Hoffmann-La Roche Ltd and its affiliated company Genentech, Inc.; Fujirebio; GE Healthcare; IXICO Ltd.; Janssen Alzheimer Immunotherapy Research & Development, LLC.; Johnson & Johnson Pharmaceutical Research & Development LLC.; Lumosity; Lundbeck; Merck & Co., Inc.; Meso Scale Diagnostics, LLC.; NeuroRx Research; Neurotrack Technologies; Novartis Pharmaceuticals Corporation; Pfizer Inc.; Piramal Imaging; Servier; Takeda Pharmaceutical Company; and Transition Therapeutics. The Canadian Institutes of Health Research is providing funds to support ADNI clinical sites in Canada. Private sector contributions are facilitated by the Foundation for the National Institutes of Health (www.fnih.org). The grantee organization is the Northern California Institute for Research and Education, and the study is coordinated by the Alzheimer’s Therapeutic Research Institute at the University of Southern California. ADNI data are disseminated by the Laboratory for Neuro Imaging at the University of Southern California.

## References

1. WHO. Public health response to dementia [Internet]. Geneva: World Health Organization. 2021. 137 p. Available from: https://www.who.int/publications/i/item/9789240033245

2. Sudre CH, Cardoso MJ, Modat M, Ourselin S. Imaging biomarkers in Alzheimer’s disease. In: Handbook of Medical Image Computing and Computer Assisted Intervention [Internet]. Elsevier; 2020. p. 343–78. Available from: https://linkinghub.elsevier.com/retrieve/pii/B978012816176000020X

3. Lin SY, Lin PC, Lin YC, Lee YJ, Wang CY, Peng SW, et al. The Clinical Course of Early and Late Mild Cognitive Impairment. Front Neurol. 2022;13(May):1–10.

4. Atalar A. Early detection of alzheimer’s disease with deep learning using 3d MRI and patient informations. 2022;

5. Yang J, Feng X, Laine AF, Angelini ED. Characterizing Alzheimer’s Disease with Image and Genetic Biomarkers Using Supervised Topic Models. IEEE J Biomed Heal Informatics. 2020;24(4):1180–7.

6. Okyay S, Adar N. Dementia-related user-based collaborative filtering for imputing missing data and generating a reliability scale on clinical test scores. PeerJ. 2022;10:1–19.

7. Liu M, Zhang J, Adeli E, Shen Di. Joint classification and regression via deep multi-task multi-channel learning for Alzheimer’s disease diagnosis. IEEE Trans Biomed Eng. 2019;66(5):1195–206.

8. Pillai PS, Leong TY. Fusing Heterogeneous Data for Alzheimer’s Disease Classification. Stud Health Technol Inform. 2015;216:731–5.

9. Kaur J, Shekhar C. Multimodal medical image fusion using deep learning. In: Advances in Computational Techniques for Biomedical Image Analysis [Internet]. Elsevier; 2020. p. 35–56. Available from: https://linkinghub.elsevier.com/retrieve/pii/B9780128200247000025

10. Sørensen L, Nielsen M. Ensemble support vector machine classification of dementia using structural MRI and mini-mental state examination. J Neurosci Methods [Internet]. 2018 May;302:66–74. Available from: https://linkinghub.elsevier.com/retrieve/pii/S0165027018300177

11. Forouzannezhad P, Abbaspour A, Li C, Cabrerizo M, Adjouadi M. A Deep Neural Network Approach for Early Diagnosis of Mild Cognitive Impairment Using Multiple Features. Proc -17th IEEE Int Conf Mach Learn Appl ICMLA 2018. 2018;1341–6.

12. Liu S, Liu S, Cai W, Che H, Pujol S, Kikinis R, et al. Multimodal Neuroimaging Feature Learning for Multiclass Diagnosis of Alzheimer’s Disease. IEEE Trans Biomed Eng. 2015;62(4):1132–40.

13. Punjabi A, Martersteck A, Wang Y, Parrish TB, Katsaggelos AK. Neuroimaging modality fusion in Alzheimer’s classification using convolutional neural networks. PLoS One [Internet]. 2019;14(12):1–14. Available from: 10.1371/journal.pone.0225759

14. Zhang Y, Wang S, Xia K, Jiang Y, Qian P. Alzheimer’s disease multiclass diagnosis via multimodal neuroimaging embedding feature selection and fusion. Inf Fusion [Internet]. 2021 Feb;66:170–83. Available from: https://linkinghub.elsevier.com/retrieve/pii/S1566253520303638

15. Lin W, Gao Q, Du M, Chen W, Tong T. Multiclass diagnosis of stages of Alzheimer’s disease using linear discriminant analysis scoring for multimodal data. Comput Biol Med [Internet]. 2021 Jul;134:104478. Available from: https://linkinghub.elsevier.com/retrieve/pii/S0010482521002729

16. Pelka O, Friedrich CM, Nensa F, Mönninghoff C, Bloch L, Jöckel KH, et al. Sociodemographic data and APOE-ε4 augmentation for MRI-based detection of amnestic mild cognitive impairment using deep learning systems. PLoS One. 2020;15(9 September):1–24.

17. Payan A, Montana G. Predicting Alzheimer’s disease a neuroimaging study with 3D convolutional neural networks. ICPRAM 2015 - 4th Int Conf Pattern Recognit Appl Methods, Proc. 2015;2:355–62.

18. 18. Hosseini-Asl E, Keynton R, El-Baz A. Alzheimer’s disease diagnostics by adaptation of 3D convolutional network. Proc - Int Conf Image Process ICIP. 2016;2016-Augus(502):126– 30.

19. Sahumbaiev I, Popov A, Ramírez J, Górriz JM, Ortiz A. 3D-CNN HadNet classification of MRI for Alzheimer’s Disease diagnosis. 2018 IEEE Nucl Sci Symp Med Imaging Conf NSS/MIC 2018 - Proc. 2018;3–6.

20. Helaly HA, Badawy M, Haikal AY. Deep Learning Approach for Early Detection of Alzheimer’s Disease. Cognit Comput [Internet]. 2022;14(5):1711–27. Available from: 10.1007/s12559-021-09946-2

21. Jayanthi VS, Simon BC, Baskar D. Alzheimer’s disease classification using deep learning. In: Computational Intelligence and Its Applications in Healthcare [Internet]. Elsevier; 2020. p. 157–73. Available from: https://linkinghub.elsevier.com/retrieve/pii/B978012820604100011X

